# Genetic evidence for the beneficial effect of greater fat-free mass on cardiometabolic health

**DOI:** 10.64898/2025.12.02.25341439

**Authors:** Xuye Jiang, Mario García-Ureña, César Cunha, María José Romero-Lado, Thorkild I.A. Sørensen, Tuomas O. Kilpeläinen

## Abstract

**Background:** Fat-free mass (FFM), particularly skeletal muscle, is widely considered beneficial for glucose homeostasis and cardiometabolic health. However, its independent role remains uncertain due to confounding by other cardiometabolically relevant anthropometric traits, namely fat mass (FM), fat distribution, and height. Genetic approaches offer a unique opportunity to disentangle these effects and minimize bias.

**Aim:** We aimed to assess the independent effects of FFM on glucose and insulin homeostasis, type 2 diabetes (T2D) risk, and cardiovascular risk factors and events using two complementary genetic approaches.

**Methods:** We first applied multivariable Mendelian randomization (MVMR) to estimate the effect of FFM while accounting for FM, fat distribution, and height. Second, we developed an FFM-specific polygenic score by excluding variants associated with these traits. Genetic instruments for FFM were derived from 1,209 independent genome-wide significant variants among 361,918 UK Biobank participants with bioimpedance data. The polygenic score, comprising 58 of these variants, was validated in an independent sample of 34,540 individuals with DXA data. Cardiometabolic outcome effects were tested using summary statistics from the largest available GWAS meta-analyses and the FinnGen study (combined *n* up to 1.8 million).

**Results:** Both MVMR and the FFM-specific polygenic score indicated a protective effect of higher FFM on postprandial glucose tolerance, insulin sensitivity, and T2D risk (MVMR: OR=0.83, 95%CI 0.76-0.91; Polygenic score: OR=0.81, 95%CI 0.74-0.88 per SD increase in FFM). They also indicated a beneficial effect on cardiometabolic risk markers and risk of stroke (MVMR: OR=0.84, 95%CI 0.77-0.92; Polygenic score: OR=0.79, 95%CI 0.71-0.89), with a more modest reduction observed for myocardial infarction (MVMR: OR=0.89, 95%CI 0.81-0.98; Polygenic score: OR=0.90, 95%CI 0.81–1.01).

**Conclusion:** Genetic evidence supports a beneficial effect of greater FFM on cardiometabolic health, underscoring the value of preserving or increasing FFM in T2D and CVD prevention.

## INTRODUCTION

Obesity is a well-established risk factor for type 2 diabetes (T2D) and cardiovascular disease (CVD)^1^, and is conventionally assessed using body mass index (BMI). However, as a metric based on weight and height, BMI fails to distinguish between fat mass (FM) and fat-free mass (FFM), thereby obscuring their distinct metabolic roles. This distinction is important as FFM, particularly skeletal muscle, has been associated with favorable cardiometabolic outcomes, such as enhanced glucose homeostasis^2^, increased resting metabolic rate^3^, reduced blood pressure^4,5^, and lower all-cause mortality^6^.

Despite its potential protective effects, the independent contribution of FFM to cardiometabolic health remains unclear due to its strong and time-varying correlation with other cardiometabolically relevant anthropometric traits – FM, fat distribution, and height – making it difficult to isolate the specific effects of FFM^7^. Disentangling these relationships is essential for guiding effective prevention and treatment strategies for T2D and CVD. For example, weight loss interventions that inadvertently reduce FFM may compromise metabolic health, whereas strategies that preserve or increase FFM – such as resistance training – could offer additional benefits ^8^.

The emergence of novel pharmacological treatments for obesity, particularly GLP-1 receptor agonists and dual agonists, has transformed the landscape of weight management and cardiometabolic risk reduction ^9^. While these therapies have demonstrated substantial benefits in reducing fat mass and improving glycemic control, recent evidence has raised concerns about their unintended effects on FFM, including skeletal muscle loss^8^. Given the central role of FFM in cardiometabolic health, understanding its independent contribution to disease risk is increasingly important for evaluating the long-term metabolic consequences of these interventions.

Genetic approaches offer a powerful tool to address these challenges. Since genetic variants are randomly allocated at conception and remain fixed throughout life, they can serve as proxies for modifiable exposures, providing an opportunity to investigate their causal effects on disease risk. Prior studies using Mendelian randomization have suggested that FFM may reduce the risk of T2D, myocardial infarction, and stroke^10^. However, these studies relied on FFM and FM indexed to height-squared and did not account for effects on body fat distribution, leaving room for residual confounding.

In the present study, we overcome these limitations by employing two complementary genetic approaches. First, we use multivariable Mendelian randomization (MVMR) to estimate the independent causal effects of FFM on glucose and insulin homeostasis, cardiovascular risk factors, and risks of T2D, myocardial infarction, and stroke, while adjusting for FM, fat distribution (waist-hip ratio, WHR), and height. Second, we develop and validate a polygenic score specific to FFM by excluding variants associated with FM, fat distribution, or height, to further isolate the role of FFM in cardiometabolic health. By integrating evidence from both approaches, we provide more robust evidence on the beneficial effect of FFM on cardiometabolic health and its potential as a modifiable target for the prevention of T2D and CVD.

## METHODS

### Study Design

We investigated the independent effects of FFM on cardiometabolic health using two complementary genetic approaches: (i) MVMR analysis to estimate the effect of FFM while adjusting for FM, body fat distribution, and height; and (ii) Development and validation of an FFM-specific polygenic score, constructed by excluding variants associated with FM, fat distribution, or height. First, we conducted a GWAS to identify loci associated with FFM using a UK Biobank sample comprising 361,918 individuals (mean age 56.7 years, 56% women) of European ancestry whose body composition was assessed by bioimpedance (**Supplementary Table 1**). We identified 1,209 independent (r^2^<0.01 within ±500 kb from the lead variant), genome-wide significant (P<5x10^-8^) variants, which we subsequently used as (i) instruments in MVMR analyses, and (ii) to select variants for an FFM-specific polygenic score. We independently validated the FFM-specific polygenic score in a UK Biobank sample of 34,540 individuals of European ancestry (mean age 55.3 years, 52% women) whose body composition was assessed by dual-energy X-ray absorptiometry (DXA) (**Supplementary Table 2**). Following validation, we applied the score to largest available GWAS meta-analysis summary statistics and the FinnGen study to evaluate its associations with cardiometabolic outcomes.

### Participants

We utilized individual-level data from the UK Biobank to perform a GWAS for FFM, identify genome-wide significant variants, and develop and validate an FFM-specific polygenic score. In addition, the UK Biobank data were used to assess the generalizability of the polygenic score across diverse ancestries and compare its cardiometabolic effects between sexes.

The UK Biobank is a prospective cohort study conducted across 22 study centers in the United Kingdom, which recruited over 500,000 participants aged 40 to 69 years between 2006–2010^11^. During the baseline study visit, participants completed a touchscreen questionnaire, underwent a nurse-led interview covering lifestyle and medical history, and provided physical measurements and biological samples.

From the total cohort 502,365 participants, our GWAS for FFM excluded 77,640 individuals who were of non-European origin based on either genetic data or self-report, 6,617 lacking body composition measurement from either bioimpedance or DXA, 19,351 diagnosed with myocardial infarction or T2D by the recruitment date, 945 who withdrew consent during follow-up, 325 with discordant genetic or self-reported sex, 417 with sex chromosome aneuploidy, 444 identified as outliers in heterozygosity or missingness, and 169 with ten or more third-degree relatives in the study sample. After exclusions, 396,458 individuals of European ancestry remained for analysis (**Supplementary Figure 1**). To ensure unbiased validation of the polygenic score, we divided this sample into a discovery cohort (those without DXA measurements) and a validation cohort (34,540 individuals with DXA measurements). The discovery cohort was used for the GWAS and derivation of the FFM-specific polygenic score, while the validation cohort provided an independent dataset for evaluation of the score. In follow-up analyses, we also included 10,603 UK Biobank participants of Asian ancestry and 7,368 of African ancestry, to evaluate ancestry-specific effects.

In parallel, we analyzed summary-level GWAS data from the FinnGen cohort, a nationwide Finnish biobank that integrates genomic data with longitudinal health registry information^12^. The cohort comprises approximately 217,000 men (43%) and 281,000 women (57%), with a median age of 53 years at sample collection.

Lastly, we incorporated summary GWAS meta-analysis results for each cardiometabolic outcome, with study-specific details provided in **Supplementary Table 3.**

### Genotyping in the UK Biobank

Genome-wide genotypes were available for approximately 480,000 participants of the UK Biobank. Genotyping was performed using two closely related purpose-designed arrays: the UK BiLEVE Axiom array for ∼50,000 participants and the UK Biobank Axiom array for the remaining ∼430,000 participants. The genotypes were imputed from the Haplotype Reference Consortium and UK10K haplotype reference panels.

### Phenotyping in the UK Biobank

In the UK Biobank, baseline age and sex were recorded via a touchscreen questionnaire. Body weight, height, waist circumference, hip circumference, blood pressure, and body composition (Tanita BC418MA Body Composition Analyzer) were measured for all participants at baseline. Some participants underwent additional body imaging measurements by DXA scans^13^. These data were available for 34,540 individuals at the time of our analysis. Blood glucose, HbA1c, triglycerides (TG), total cholesterol, LDL cholesterol, HDL cholesterol, and C-reactive protein were measured from blood samples collected at recruitment. The participants were not requested to fast overnight before blood sampling.

Incident cases of T2D in the UK Biobank were identified using the ICD-10 code E11 (non-insulin dependent diabetes mellitus) derived from linked primary care records, hospital inpatient records, self-reported diagnoses, and death registry information. Incident myocardial infarction and stroke were identified using algorithmically defined outcomes curated by the UK Biobank Outcome Adjudication Group, which integrates clinical codes across multiple data sources to enhance diagnostic accuracy^14^. Time-to-event was calculated from the date of baseline assessment to the earliest occurrence of incident T2D, myocardial infarction, stroke, death, or censoring at the end of follow-up. Follow-up concluded on September 1, 2023 for myocardial infarction and stroke, and on October 1, 2023 for T2D. Participants with prevalent T2D, myocardial infarction, or stroke at baseline were excluded. To minimize misclassification, individuals without a formal T2D diagnosis but random glucose above 11.1 mmol/L or HbA1c above 6.5% at baseline were also excluded.

### Identification of FFM-associated genetic variants

To identify genetic variants associated with FFM, we performed GWAS in our UK Biobank discovery cohort, including 361,918 European-ancestry individuals with bioimpedance-derived body composition measures. GWAS analyses were performed for FFM, FM, height, and WHR, adjusting for age, age², sex, genotyping array, center, and the first 15 genetic principal components. For WHR, we further adjusted for FM to isolate independent genetic effects. We normalized the distributions of FFM, FM, WHR adjusted for FM (WHR_adjFM_), and height using rank-based inverse normal transformation, applied separately for women and men, to account for sex-specific distributional differences. GWAS were implemented using the REGENIE tool^15^, which involves two analysis steps. In the first step, a subset of genetic markers is used to fit a whole-genome regression model that captures the fraction of phenotype variance attributable to genetic effects. Variants were excluded from this step if they had a minor allele frequency ≤ 0.001, minor allele count ≤ 100, genotype missingness ≥ 10%, or Hardy-Weinberg equilibrium p-value ≤ 10^−1^^5^. In the second step, REGENIE tests each genetic marker for association with the phenotype conditional on the prediction from the first step, using a leave-one-chromosome-out scheme. In this step, we additionally excluded variants with imputation quality scores ≤ 0.3, multiallelic variants, and ambiguous SNPs. To evaluate GWAS quality, we applied linkage disequilibrium score (LDSC) regression (**Supplementary Figures 2** **and 3**). The resulting attenuation ratio (<0.08) and an LDSC intercept (<1.5), indicated adequate control for inflation and genetic confounding^16^ (**Supplementary Figures 2** **and 3**).

### Multivariable Mendelian randomization analysis

To estimate the independent causal effects of FFM on cardiometabolic events and risk factors, we performed MVMR using genome-wide significant (P<5x10^-8^), independent (r^2^<0.01 within ±500 Kb) SNPs associated with FFM in our UK Biobank discovery sample. These SNPs served as genetic instruments in MVMR models that accounted for FM, WHR_adjFM_, and height as covariates (**Supplementary Table 4**). MVMR analyses were conducted using MR-Horse, a Bayesian approach designed to maintain low type I error rates under conditions of high-pleiotropy and weak instrument strength^17^.

Effect estimates for cardiometabolic outcomes were obtained from the largest published GWAS meta-analyses (**Supplementary Table 3**). For binary outcomes, including T2D, myocardial infarction, and stroke, we additionally performed MVMR analyses using independent summary statistics from the FinnGen cohort. These results were subsequently meta-analyzed with those from the published GWAS to enhance statistical power.

### Development and validation of an FFM-specific polygenic score

To develop an FFM-specific polygenic score, we selected all independent, genome-wide significant variants associated with FFM in our discovery cohort that were not associated with FM or WHR_adjFM_ (P>0.05) and had no primary association with height, defined as a stronger P value for height than for FFM in a matched sample size. This filtering ensured that selected variants captured FFM-specific genetic effects while minimizing confounding from other anthropometric traits.

Polygenic scores were calculated using the standard weighted sum approach^18^:

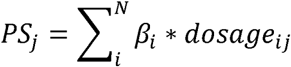

where N is the number of SNPs in the PS, β*_i_* is the effect size of SNP *i* from the FFM GWAS, and *dosage_ij_*is the number of effect alleles carried by individual *j*. All effect sizes and alleles were aligned to reflect the FFM-increasing direction.

We validated the polygenic score in an independent validation UK Biobank sample of 34,540 individuals of European ancestry whose body composition was measured by DXA and who were not included in the GWAS discovery cohort. Linear regression models were used to assess the association between the polygenic score and DXA-derived body composition measures, adjusting for age, sex, study center, and the first 15 genetic principal components. The proportion of trait variance explained by the score was quantified as the difference in R^2^ between models with and without the polygenic score^19^. To examine potential sex differences in genetic effects, we conducted sex-stratified analyses and tested for sex-by-score interaction.

### Generalizability of the FFM-specific polygenic score in Asian and African ancestries

To assess the generalizability of the FFM-specific polygenic score across diverse populations, we performed separate analyses among UK Biobank participants of Asian and African ancestry. The Asian ancestry group included individuals of Indian, Pakistani, Bangladeshi, Chinese, and other Asian backgrounds, while the African ancestry group included participants with Afro-Caribbean and other African descent. Body composition in both groups was assessed using bioimpedance. We applied the same quality control procedures to these ancestry groups as those used for European participants. After exclusions, the final analysis included 10,603 individuals of Asian ancestry and 7,368 individuals of African ancestry. In each ancestry group, we derived both weighted and unweighted versions of the FFM polygenic score using the same 58 genetic variants identified in the European discovery cohort. The weighted score was calculated using effect size estimates from the European GWAS, while the unweighted score assigned a fixed weight of 1 to each variant.

### Association of the FFM-specific polygenic score with cardiometabolic outcomes

We examined the associations of the FFM-specific polygenic score with cardiometabolic outcomes using summary statistics from the largest available European-ancestry GWAS for each trait, excluding UK Biobank data where possible to avoid sample overlap (**Supplementary Table 3**). Due to limited availability of sex-stratified GWAS summary data for most outcome traits, analyses were performed in a sex-combined manner. We used the grs.summary function from the gtx.package in R to calculate the weighted mean effect of the independent alleles on the risk of T2D^20^, myocardial infarction^21^, stroke^22^. The same function was applied to investigate associations with intermediate cardiometabolic traits, including fasting and 2-hour glucose^23^, HbA1c^24^, fasting insulin^24^, insulin fold change^23^, insulin sensitivity index^23^, total^25^, LDL^25^ and HDL cholesterol^25^, TG ^25^, TG/HDL cholesterol ratio, C-reactive protein^26^, and systolic and diastolic blood pressure^27^.

To further evaluate the association of the FFM-specific polygenic score with incident T2D and CVD outcomes, and to explore potential sex differences, we applied Cox proportional hazards regression analyses in the UK Biobank, stratified by sex. For quantitative traits, we applied linear regression models on randomly measured glucose, HbA1c, total cholesterol, LDL cholesterol, HDL cholesterol, TG, TG/HDL cholesterol ratio, C-reactive protein, and systolic and diastolic blood pressure. All models were adjusted for age, study center, and the first 15 genetic principal components. Quantitative outcome traits were rank inverse normal transformed prior to analysis to ensure normality (**Supplementary Figure 4**).

To facilitate interpretation, effect estimates for cardiometabolic outcomes are reported per standard deviation increase (SD) in FFM (equivalent to 11.45 kg) predicted by the polygenic score in the UK Biobank.

## RESULTS

### Assessing the effects of FFM on cardiometabolic health using MVMR

We performed MVMR analyses using 1,209 independent genome-wide significant variants associated with FFM to assess its independent effects on cardiometabolic outcomes while accounting for FM, WHR_adjFM_, and height. We further incorporated independent summary results from the FinnGen study and combined them with the GWAS meta-analysis results using fixed effects meta-analysis to enhance statistical power.

The MVMR analysis indicated that higher FFM was associated with improved postprandial glucose tolerance and insulin action, as evidenced by lower 2-hour glucose levels and insulin fold change, as well as higher insulin sensitivity index during an oral glucose tolerance test. Furthermore, TG/HDL-C ratio, a surrogate marker of insulin resistance, was reduced **(Figure 1 A)**. In contrast, no significant associations were observed between FFM and fasting glucose, fasting insulin, or HbA1c levels. Higher FFM was also associated with favorable profiles of several cardiovascular risk markers, including total and HDL and LDL cholesterol, TG, C-reactive protein (**Figure 1 A**), and systolic and diastolic blood pressure (**Figure 1 B**).

**Figure 1.**
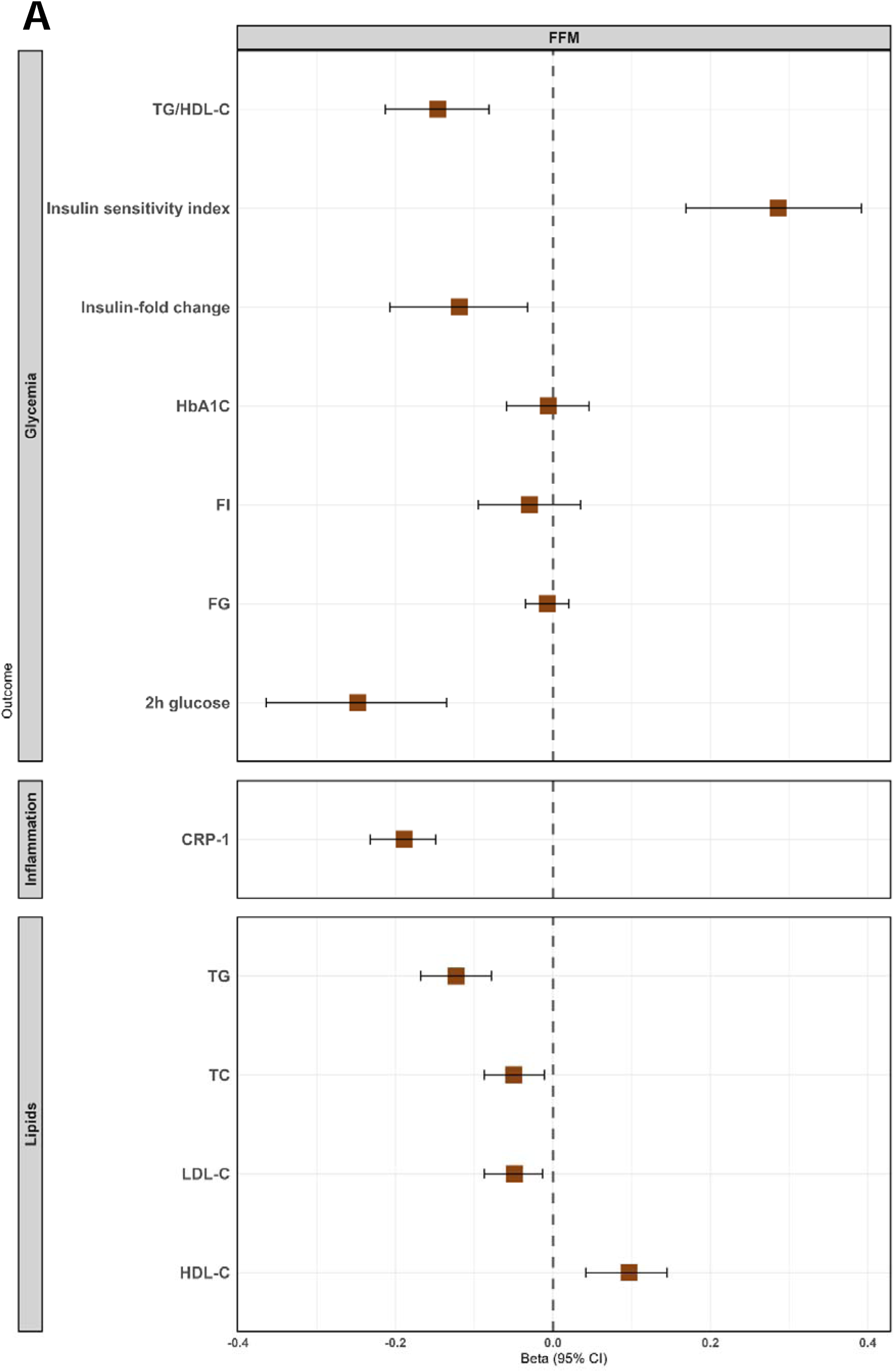
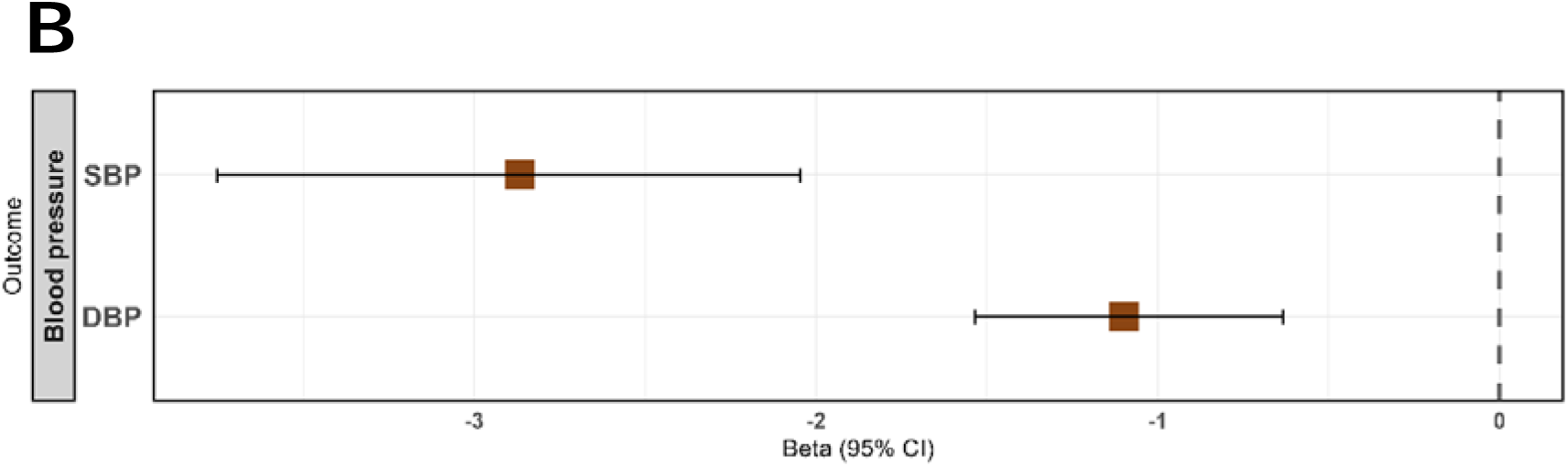
Association of FFM with Cardiometabolic Risk factors Accounting for Fat Mass, Height and WHRajdFM using multivariable Mendelian randomization analyses The forest plot displays the association of fat-free mass (FFM) with cardiometabolic risk factors, adjusted for fat mass (FM), height, and waist-to-hip ratio adjusted for fat mass (WHRajdFM) using multivariable Mendelian randomization analyses. The statistical significance threshold for analyses reported in this panel was P < 0.05.

Regarding disease outcomes, increased FFM had a protective effect on risk of T2D in both the GWAS meta-analysis (OR=0.80, 95%CI 0.70-0.91) and FinnGen (OR=0.87, 95%CI 0.76-0.99), and the combined meta-analysis corroborated this protective effect (OR=0.83, 95%CI 0.76–0.91, P=1.0x10^⁻6^) (**Figure 2**). For myocardial infarction, the effect was non-significant in the GWAS meta-analysis (OR=0.88, 95%CI 0.78–1.00) or FinnGen (OR=0.91, 95%CI 0.78–1.06), but resulting in a combined estimate that supported a protective effect (OR=0.89, 95%CI 0.81–0.98, P=0.02). Similarly, while the GWAS meta-analysis did not show a significant effect of FFM on stroke (OR=0.89, 95%CI 0.78–1.02), the FinnGen results (OR=0.81, 95%CI 0.72–0.92) and the combined estimate (OR=0.84, 95%CI 0.77–0.92, P=1.2x10^⁻4^) indicated a protective effect (**Figure 2**).

**Figure 2.**
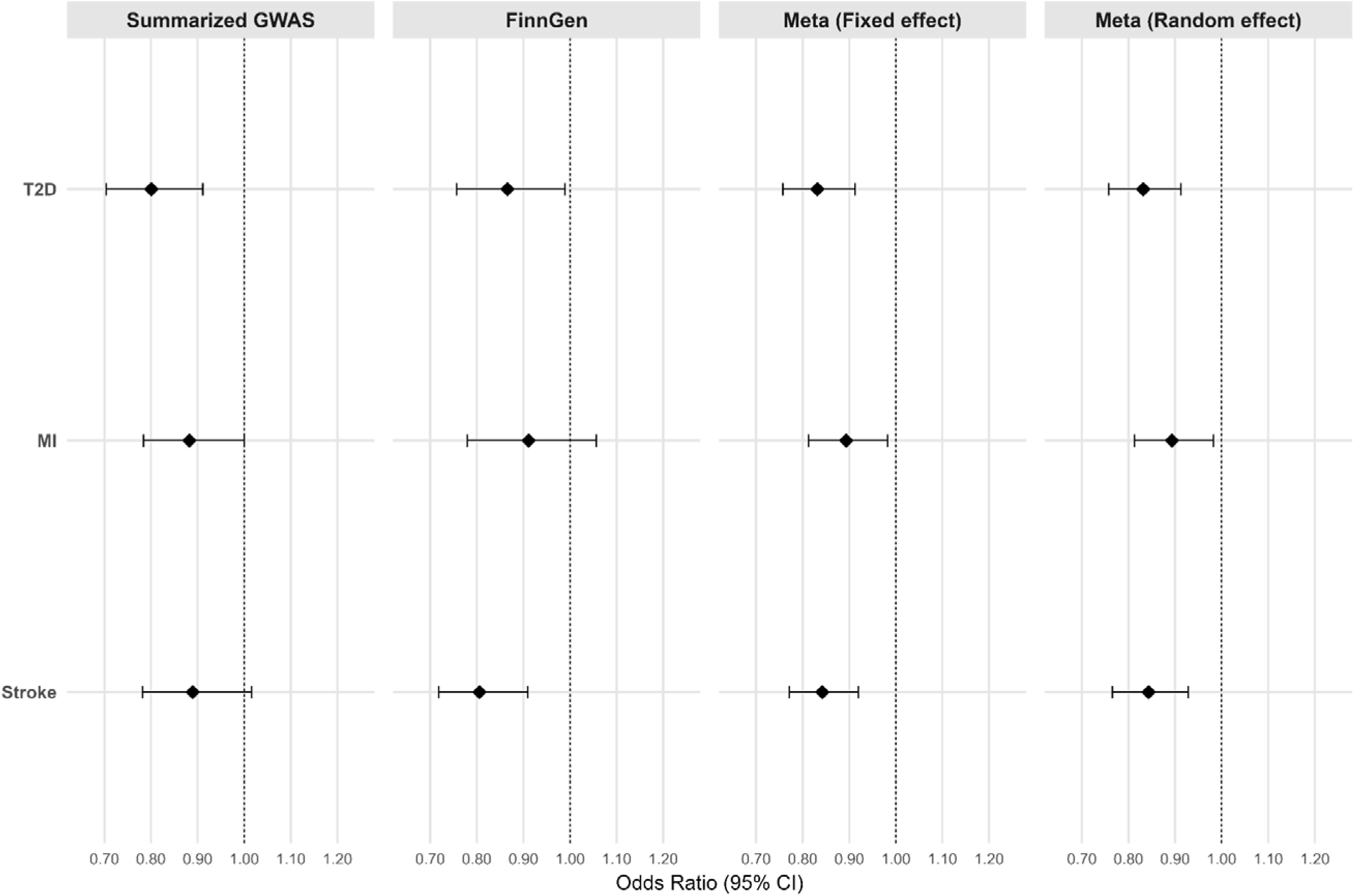
**Association of Fat-free Mass and Cardiometabolic Disease Accounting for Fat Mass, Height and WHRajdFM using multivariable Mendelian randomization analyses** The forest plot displays the association of fat-free mass (FFM) with cardiometabolic disease outcomes, adjusted for fat mass (FM), height, and waist-to-hip ratio adjusted for fat mass (WHRajdFM) using multivariable Mendelian randomization analyses. To ensure robustness, analyses were replicated using two independent data sources: 1) GWAS summary statistics from Consortium studies or previously published GWAS, and 2) FinnGen GWAS summary statistics for the same outcomes. The statistical significance threshold for analyses reported in this panel was P < 0.05.

### Development and validation of an FFM-specific polygenic score

To complement the MVMR findings and further isolate the role of FFM in cardiometabolic health, we developed a polygenic score specifically enriched for FFM-related genetic variation, excluding variants with primary associations with FM, body fat distribution, or height. To our knowledge, this is the first polygenic score constructed to capture FFM-specific genetic effects while removing confounding adiposity and height signals.

From the 1,209 independent genome-wide significant SNPs identified in the UK Biobank discovery cohort, we excluded 916 SNPs associated with FM (P<0.05), 119 associated with body fat distribution (WHR_adjFM_) (P<0.05), and 116 SNPs with stronger associations with height than with FFM (based on P-values in matched sample sizes). The remaining 58 FFM-specific variants were used to construct the polygenic score (**Supplementary Table 5**).

In an independent validation cohort of 34,540 UK Biobank participants with DXA-derived body composition measures, the FFM-specific polygenic score was strongly associated with higher FFM (P=6.7x10^-32^), but not with FM (P=0.28) or WHR_adjFM_ (P=0.21). A secondary association with height was observed (P=6.4x10^-10^), consistent with the expected contribution of bone mass – a component of FFM – to overall skeletal size and stature. However, the effect size for height (P=6.4x10^-10^) was substantially smaller than for FFM (P=6.4x10^-32^), indicating that the score predominantly captures variation in FFM rather than stature (**Supplementary Figure 5**). The association with FFM was consistent across sexes (P_sex-interaction_=0.84) and there was no association with FM in either sex (P=0.14 and P=0.81, respectively) (**Supplementary Table 6**). In sex-combined analysis, the score explained 0.5% of the variance in FFM (**Supplementary Figure 5**)

To further characterize the phenotype captured by the score, we examined its associations with lean and bone mass across body regions using DXA data. The score was not preferentially associated with either lean or bone mass and showed no significant regional differences across gynoid, android, arm, or leg compartments, suggesting that it reflects a generalized increase in FFM throughout the body rather than a compartment-specific effect (**Supplementary Table 7)**.

### Generalizability of the FFM-specific polygenic score in diverse ancestries

In participants of Asian ancestry, both the unweighted and weighted polygenic scores were significantly associated with higher FFM (P=3.6x10^-4^ and P=1.2x10^-4^, respectively) with no evidence of association with FM (P=0.66 and P=0.88, respectively) (**Supplementary Table 8**). Similarly, in participants of African ancestry, both score versions were associated with increased FFM (P=0.02 and P=0.008, respectively), and not with FM (P=0.43 and P=0.31, respectively). (**Supplementary Table 8**). These results suggest that the FFM-specific polygenic score captures biologically relevant variation across diverse ancestries, independent of FM. However, due to limited sample sizes, we were unable to robustly assess whether the score’s associations with cardiometabolic outcomes extend beyond European ancestry.

### Association of the FFM-specific polygenic score with cardiometabolic risk profile

Examining the largest available GWAS summary results for each outcome, the FFM-specific score showed a beneficial effect on glucose tolerance and insulin action, including lower 2-hour glucose levels (P=7.2x10^-4^), insulin fold change (P=9.9x10^-4^), and enhanced insulin sensitivity index (P=0.002) during an oral glucose tolerance test, but not on fasting measures of insulin, glucose, or HbA1c. The FFM-specific score also showed favorable effects on several cardiovascular risk factors, including total cholesterol (P=1.3x10^-7^), LDL cholesterol (P=0.02), TG (P=9.6x10^-13^), C-reactive protein (P=2.7x10^-7^, **Figure 3 A)** , and systolic (P=4.5x10^-15^) and diastolic (P=1.3x10^-6^) blood pressure (**Figure 3 B**).

### Association of the FFM-specific polygenic score with T2D and CVD risk

We examined the association of the FFM-specific polygenic score with risk of T2D, myocardial infarction, and stroke using summary statistics from the largest available GWAS meta-analyses, complemented by independent GWAS data from the FinnGen study. To assess potential sex-specific genetic effects, we additionally analyzed individual-level data from the UK Biobank.

The FFM-specific score was consistently associated with reduced risk of T2D in both the GWAS meta-analysis (OR=0.82, 95%CI 0.72-0.93 per SD increase) and FinnGen (OR=0.80, 95%CI 0.72-0.90), yielding a combined estimate of OR=0.81 (95%CI 0.74–0.88, P=7.3x10^-7^) (**Figure 4**). For myocardial infarction, the association was significant in FinnGen (OR=0.81, 95%CI 0.68–0.97), but not in the GWAS meta-analysis (OR=0.96, 95%CI 0.84–1.11), resulting in a combined estimate suggestive of a protective effect (OR=0.90, 95%CI 0.81–1.00). For stroke, the score was significantly associated with reduced risk in FinnGen (OR=0.76, 95%CI 0.66–0.87), and although the GWAS meta-analysis did not reach significance (OR=0.86, 95%CI 0.71–1.04), the combined estimate indicated a protective effect (OR=0.79, 95%CI 0.71–0.89, P=4.6x10⁻C) (**Figure 4**).

**Figure 3.**
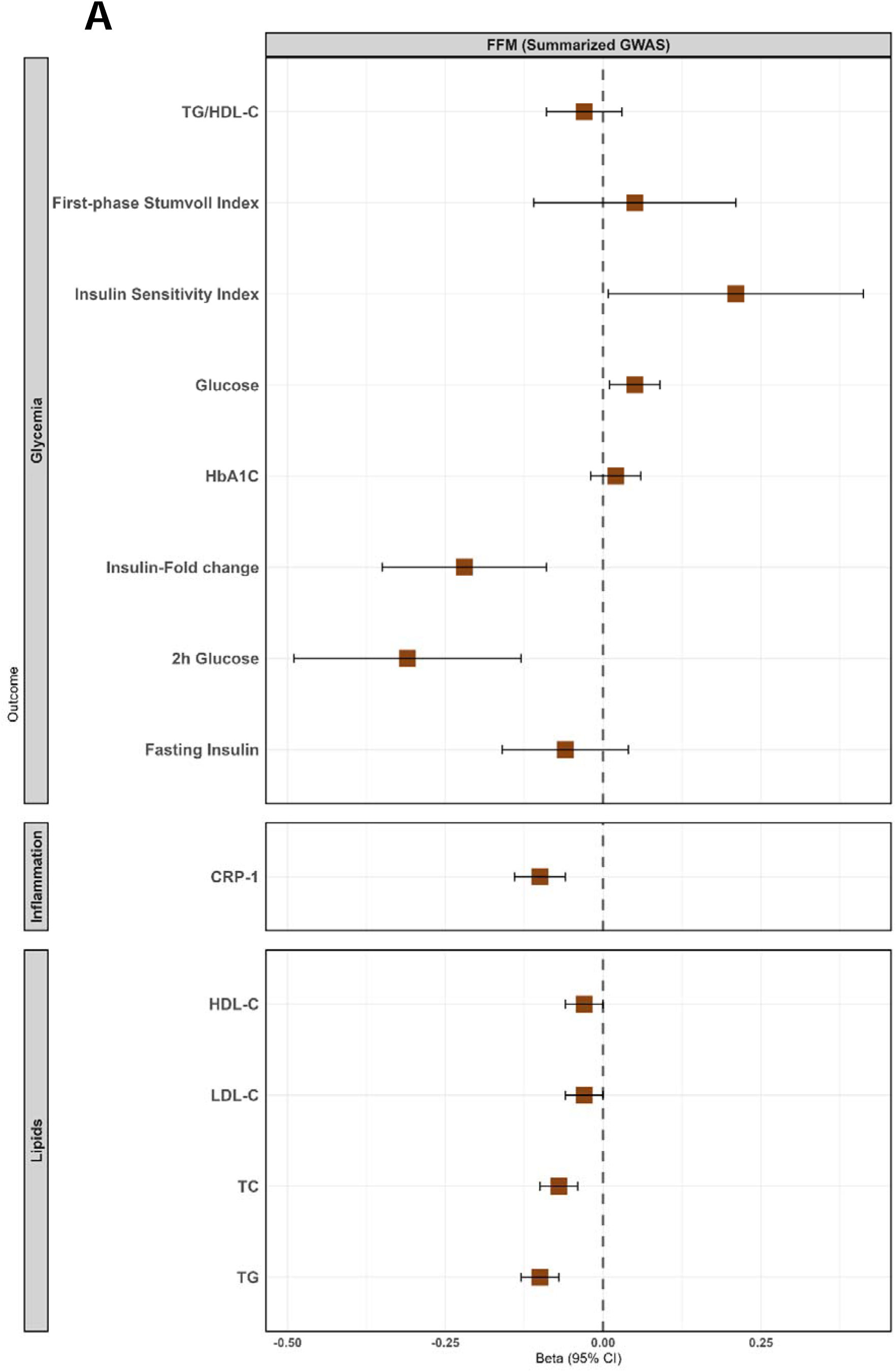
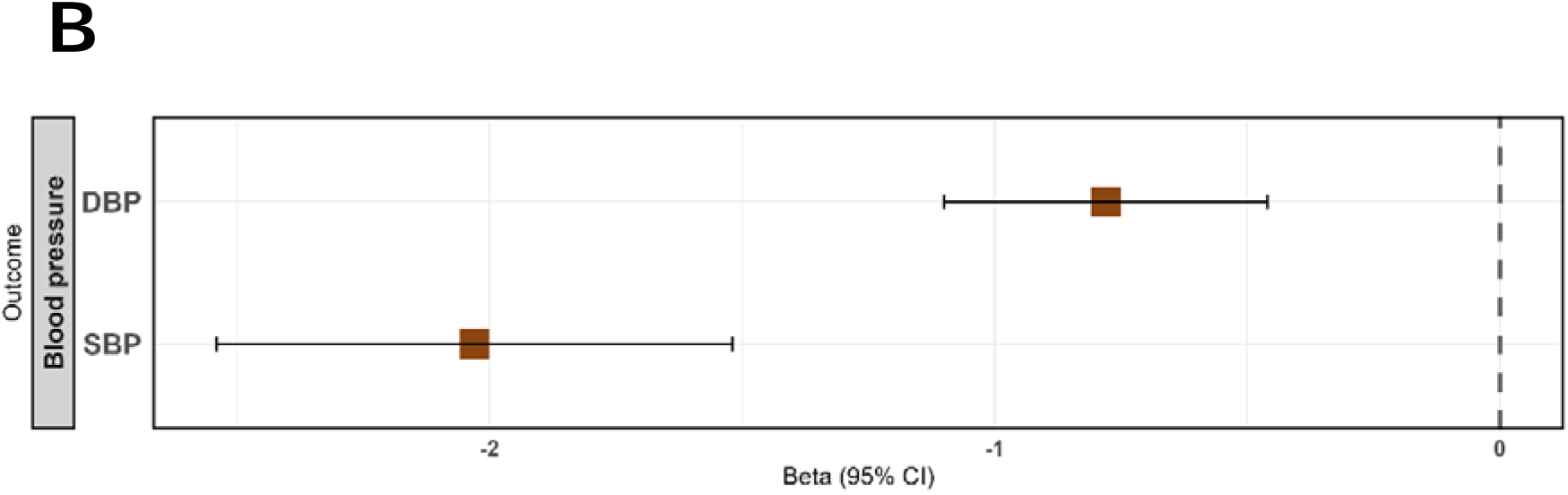
**Association with Cardiovascular Risk Factors of Fat-free Mass Specific Polygenic Score** Forest plot displays the associations of **fat-free mass (FFM)**with **cardiometabolic risk factors,** including **lipid profiles, glucose metabolism markers, blood pressure, and inflammation-related biomarkers**. Summary-level GWAS associations were estimated using the **grs.summary method,** with results reported in **per 1-SD increase in FFM (**11.45kg unit). The statistical significance threshold for analyses reported was P < 0.05.

**Figure 4.**
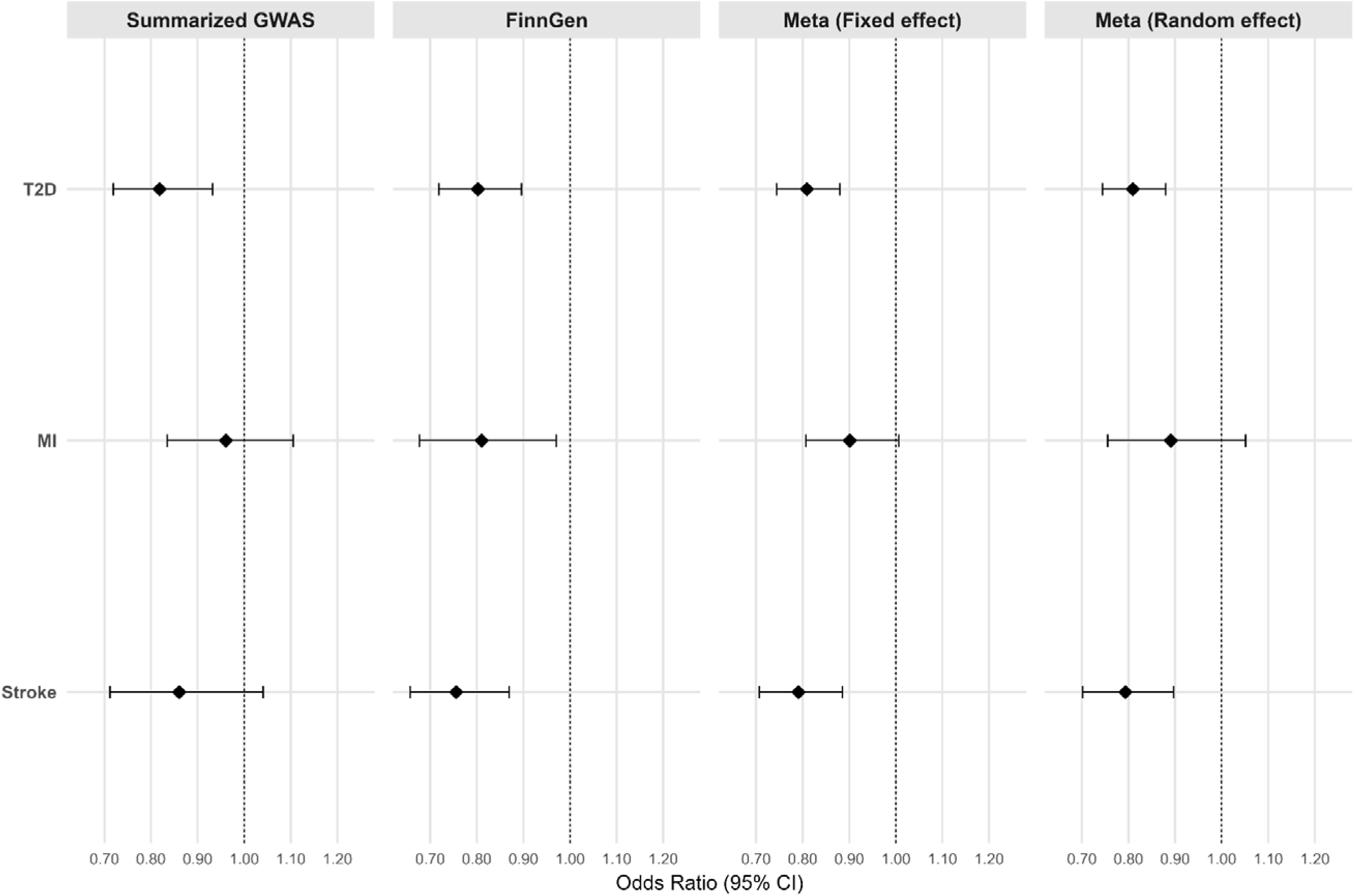
**Association with Cardiovascular Disease of Fat-free Mass Specific Polygenic Score** Forest plot displaying the associations of **fat-free mass (FFM) specific polygenic score** with **cardiometabolic diseases**, including **type 2 diabetes (T2D), myocardial infarction (MI) and stroke.** Associations were estimated using the grs.summary method and reported in odds ratio (OR) per 1-SD increase in FFM (11.45kg), with 95% confidence intervals (CIs) due to polygenic score. The statistical significance threshold for analyses reported in this panel was P < 0.05.

Sex-stratified analyses in the UK Biobank showed consistent effect sizes between women and men, with no evidence of heterogeneity (**Supplementary Table 9)**, indicating that the cardiometabolic benefits of higher genetically predicted FFM are broadly applicable across sexes.

## DISCUSSION

Although increasing FFM, particularly skeletal muscle, is widely regarded as beneficial for cardiometabolic health, empirical evidence has remained inconclusive^2^. A key challenge in observational studies is the strong correlation between FFM and other anthropometric traits – FM, fat distribution, and height – that are themselves important determinants of cardiometabolic risk, making it difficult to isolate the specific effects of FFM. To address this, we employed to complementary genetic approaches: (i) MVMR analysis, and (ii) development and validation of an FFM-specific polygenic score. Together, these strategies enabled a more precise estimation of the effects of FFM on cardiometabolic outcomes, independent of confounding traits.

Our findings indicate that higher FFM enhances glucose tolerance and insulin action, improves cardiovascular risk markers, and lowers risk T2D, stroke, and potentially myocardial infarction. These beneficial effects stand in contrast to the well-documented adverse effects of increased FM, underscoring the importance of maintaining a healthy body composition for the prevention of T2D and CVD. Our findings are particularly timely in light of the growing use of pharmacological treatments for obesity, including GLP-1 receptor agonists and dual agonists, which have demonstrated substantial efficacy in reducing FM and improving glycemic control ^9^, but also reductions in FFM, raising concerns about their long-term metabolic consequences ^6^. Our results highlight the importance of preserving or enhancing FFM during weight loss interventions, and suggest that strategies combining pharmacotherapy with resistance training or other muscle-preserving approaches may offer superior cardiometabolic benefits.

The observed associations between higher FFM and a favorable cardiovascular profile align with prior Mendelian randomization studies that used height-indexed measures of FM and FFM^28,29^. However, our study advances this literature by leveraging genetically instruments specifically enriched for FFM and by applying MVMR to account for FM, fat distribution, and height, which enabled a more targeted assessment of FFM’s cardiometabolic effects. In addition, we examined dynamic measures of glucose homeostasis during OGTT, which may better reflect skeletal muscle function than fasting measures primarily influenced by hepatic processes.

Our results suggested that the protective effect of increased FFM on T2D risk is likely mediated through enhanced glucose tolerance and insulin action, rather than changes in fasting glucose or insulin levels. This is consistent with the role of skeletal muscle as the primary site of insulin-stimulated glucose disposal via the GLUT4 transporter, which is predominantly expressed in muscle and translocates to the cell surface in response to insulin, facilitating glucose clearance from the bloodstream^30^. Greater muscle mass may amplify this mechanism, improving postprandial glucose regulation. Additional mechanisms, such as increased mitochondrial activity and the secretion of metabolically beneficial myokines, such as irisin and anti-inflammatory IL-6, may also contribute to these effects^31^.

In addition to its role in glucose regulation, increased FFM may have direct cardiovascular benefits through several mechanisms. Skeletal muscle contributes to vascular homeostasis by enhancing endothelial function and promoting vasodilation, which may lower blood pressure ^32^. Muscle tissue also facilitates lipid metabolism, enhancing the clearance of TG and LDL cholesterol ^33^, and may reduce systemic inflammation, as reflected in lower C-reative protein levels. Furthermore, the secretion of muscle-derived myokines has been implicated in vascular protection and improved metabolic signaling^31^. These mechanisms may collectively explain the observed associations between higher FFM and favorable cardiovascular risk profiles and lower risk of stroke and myocardial infarction.

Our study was limited by the focus on total FFM without accounting for cellular composition or functional quality, which may influence cardiometabolic outcomes. Moreover, while our analytical approach effectively mitigated confounding by FM, fat distribution, and height, we cannot fully exclude residual pleiotropy or interactions with other variables.

In summary, by integrating two complementary genetic strategies, we provide robust evidence that increased FFM has beneficial effects on risk of T2D, stroke, and potentially myocardial infarction, along with improvements in glucose tolerance and cardiovascular risk markers. Our findings emphasize the importance of promoting healthy body composition – particularly preserving or increasing FFM – as a potential strategy for preventing T2D and CVD.

## Data Availability

All data produced in the present study are available upon reasonable request to the authors.

## Acknowledgements

UK biobank application ID for this project is 32683. UK Biobank received ethical approval from the NorthWest Multi-Centre Research Ethics Committee, and all participants provided informed consent.

This work was supported by the Novo Nordisk Foundation (NNF22OC0074128 and NNF23SA0084103). The Novo Nordisk Foundation Center for Basic Metabolic Research (https://cbmr.ku.dk) is an independent research center at the University of Copenhagen, partially funded by an unrestricted donation from the Novo Nordisk Foundation.

M.J.R-L was supported by a research grant from the Danish Diabetes and Endocrine Academy and the Danish Cardiovascular Academy, which are funded by the Novo Nordisk Foundation, grant numbers NNF22SA0079901 and NNF20SA0067242.

---

**Supplementary Figure 1.**
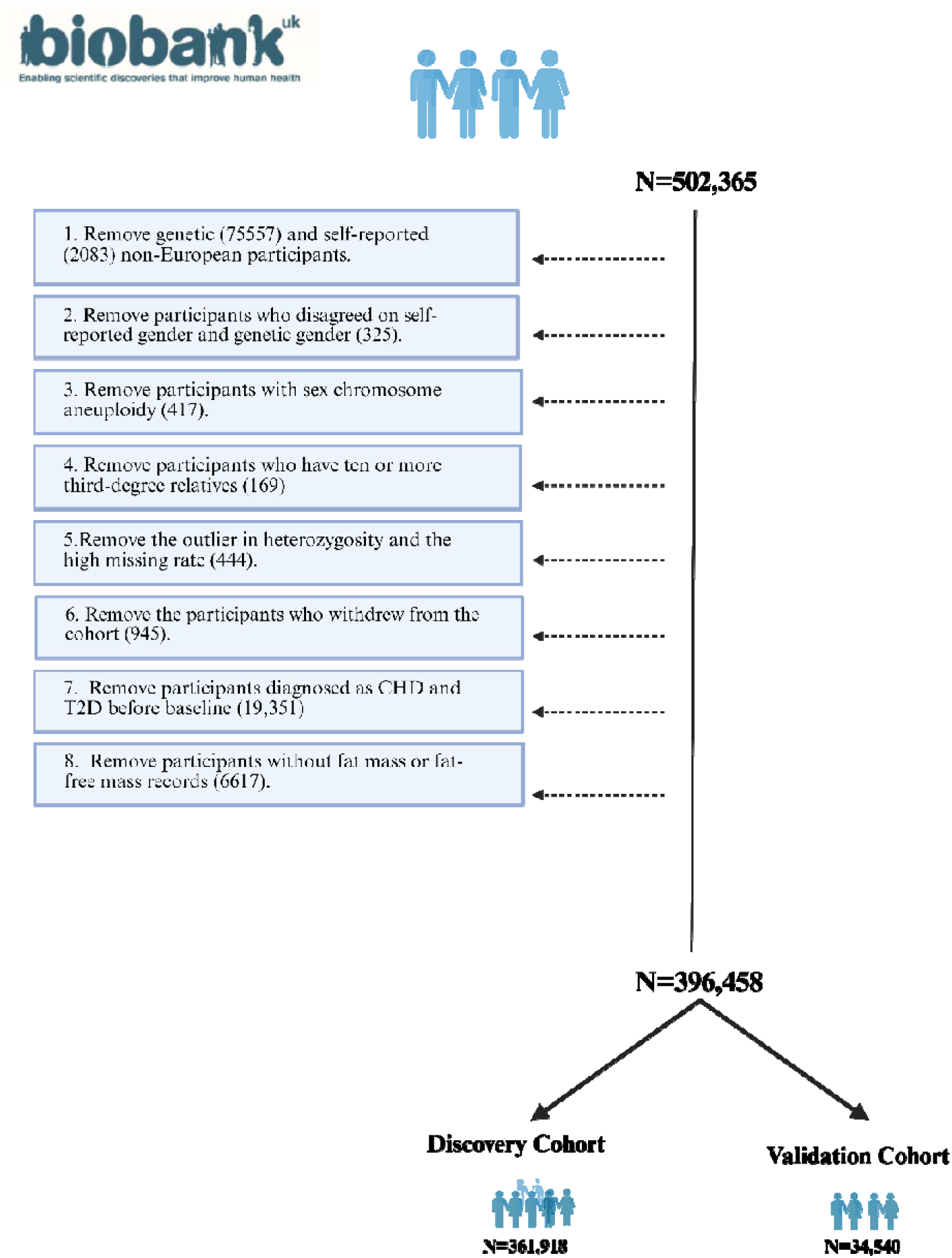
Flowchart of participant inclusion in the UK Biobank analysis. *The diagram shows the number of participants at each stage of selection, including initial recruitment, quality control exclusions, and final sample sizes used for analyses. There were no pregnant women among the participants*.

**Supplementary Figure 2.**
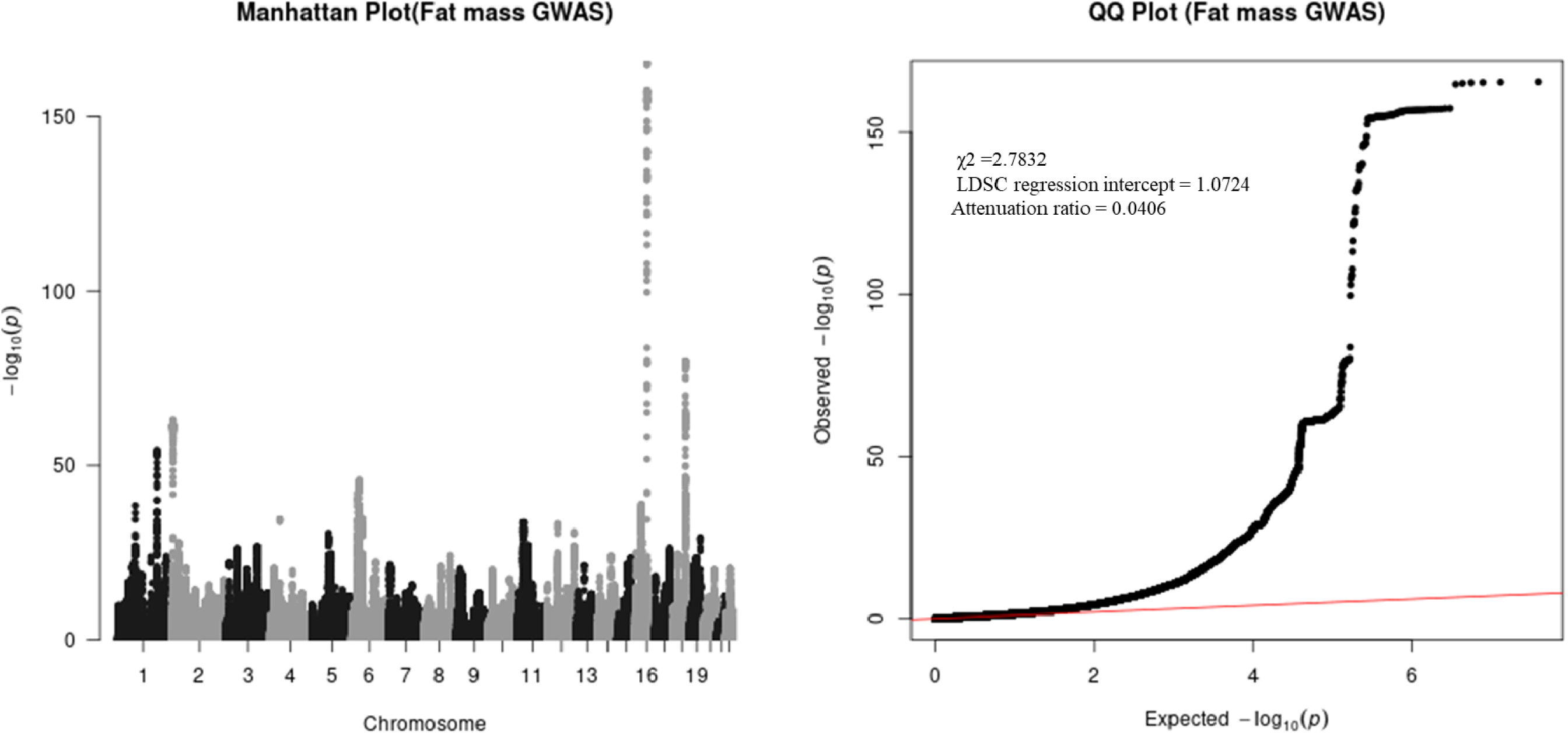
Manhattan and Quantile-Quantile Plot for the Genome-Wide Association Analysis of Fat Mass *The Manhattan plot displays –log*₁₀*(p-values) for all variants across the genome, with the genome-wide significance threshold indicated (P<5e-08). The Q-Q plot shows observed versus expected –log*₁₀*(p-values), with the mean chi-squared statistic, LDSC regression intercept, and attenuation ratio reported to assess test statistic inflation and polygenicity*.

**Supplementary Figure 3.**
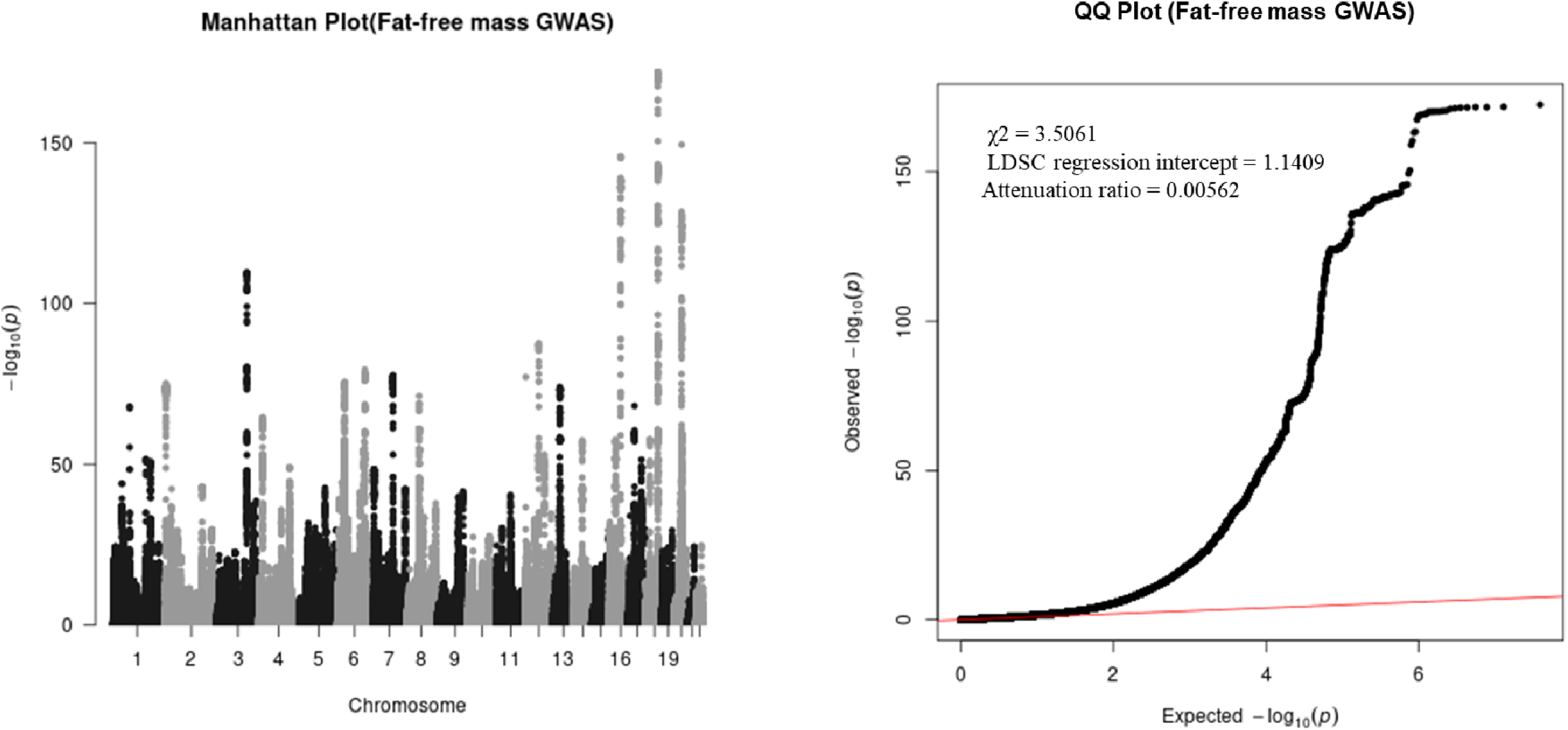
Manhattan and Quantile-Quantile Plot for the Genome-Wide Association Analysis of Fat-free Mass *The Manhattan plot displays –log*₁₀*(p-values) for all variants across the genome, with the genome-wide significance threshold indicated (P<5e-08). The Q-Q plot shows observed versus expected –log*₁₀*(p-values), with the mean chi-squared statistic, LDSC regression intercept, and attenuation ratio reported to assess test statistic inflation and polygenicity*.

**Supplementary Figure 4.**
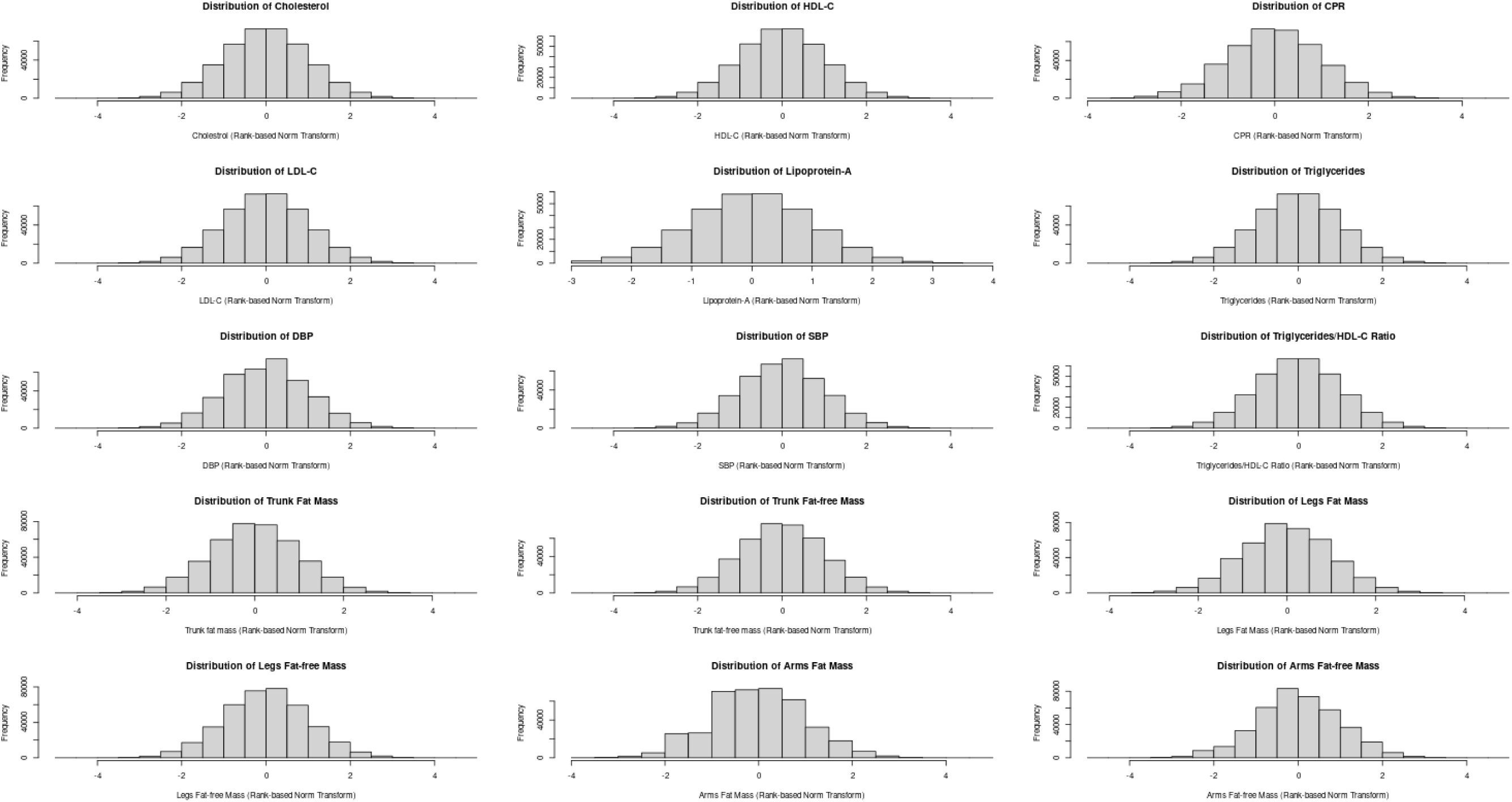
The Distribution of Continuous Outcomes *Histograms show the distributions of 15 continuous outcome variables after applying a rank-based inverse normal transformation, analyzed separately for males and females. The traits include lipid biomarkers (cholesterol, LDL-C, HDL-C, triglycerides, lipoprotein-A, and the triglycerides/HDL-C ratio), an inflammation marker (CRP), blood pressure traits (SBP and DBP), and regional body composition measures (fat mass and fat-free mass in the trunk, legs, and arms). All transformed variables approximate normal distributions*.

**Supplementary Figure 5.**
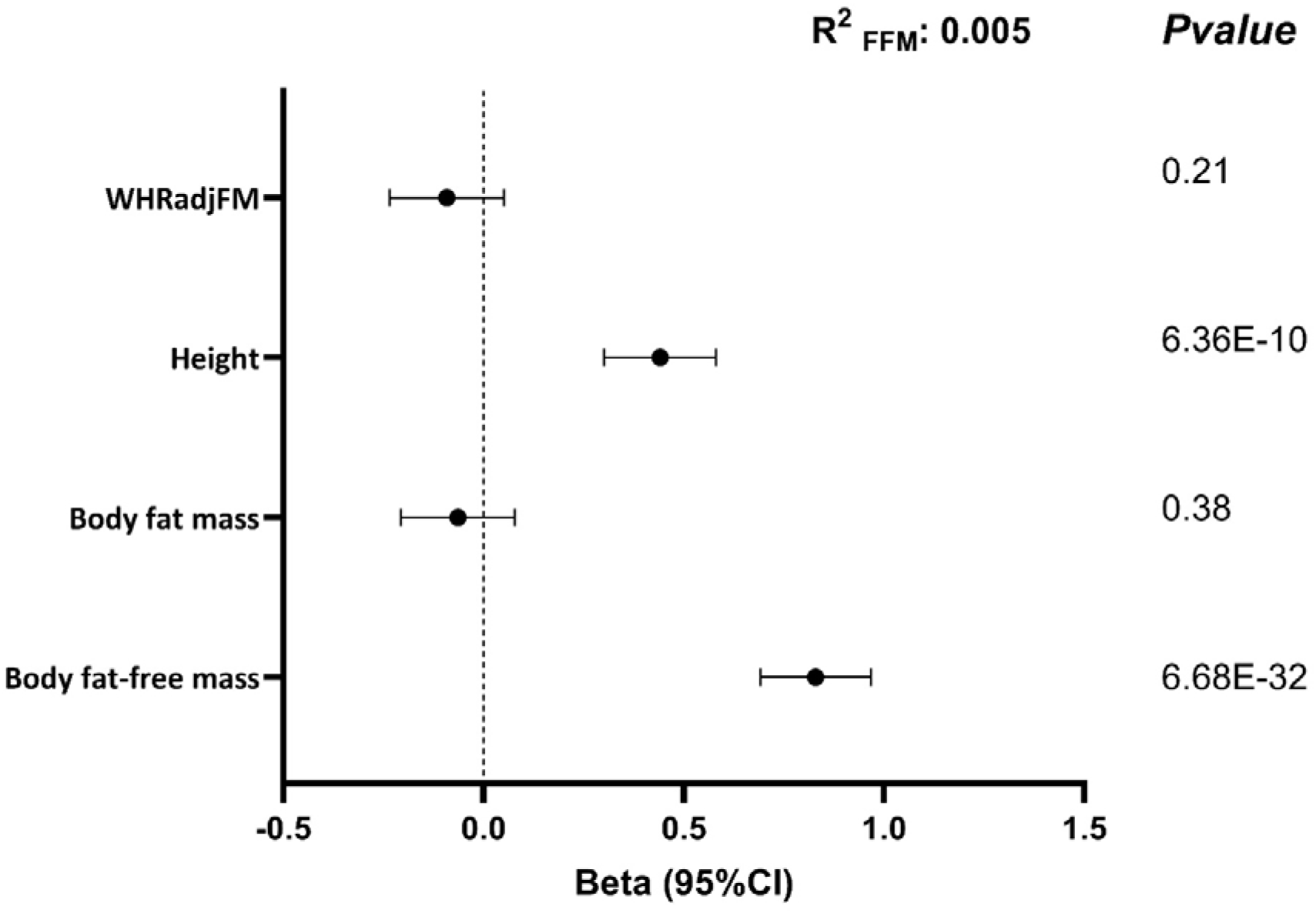
Assessment of Fat-free Mass Polygenic Score. *Forest plot displaying the associations between the fat-free mass (FFM) polygenic score and four anthropometric traits: WHRadjFM (waist-to-hip ratio adjusted for fat mass), height, body fat mass, and body fat-free mass. Associations are presented as betas per 1-SD increase in FFM (11.45 kg), with 95% confidence intervals (CIs) for each outcome. The R2 represents the proportion of variance in fat-free mass explained by the polygenic score. Linear regression models were adjusted for age, age², study center and the first 15 principal components. The statistical significance threshold for analyses reported in this panel was P < 0.05*.

